# Evolution and emergence of multidrug-resistant *Mycobacterium tuberculosis* in Chisinau, Moldova

**DOI:** 10.1101/2021.02.04.21251152

**Authors:** Tyler S. Brown, Vegard Eldholm, Ola Brynildsrud, Magnus Osnes, Natalie Stennis, James Stimson, Caroline Colijn, Sofia Alexandru, Ecaterina Noroc, Nelly Ciobanu, Valeriu Crudu, Ted Cohen, Barun Mathema

**Affiliations:** Infectious Disease Division, Massachusetts General Hospital, Boston, MA, USA; Division of Infectious Disease Control, Norwegian Institute of Public Health, Oslo, Norway; Department of Epidemiology, Mailman School of Public Health, Columbia University, New York, NY, USA; National Infection Service, Public Health England, London, UK; Department of Mathematics, Simon Fraser University, Vancouver, Canada; Phthisiopneumology Institute, Chisinau, Republic of Moldova; Department of Epidemiology (Microbial Diseases), Yale University School of Public Health, New Haven, CT, USA

**Keywords:** *Mycobacterium tuberculosis*, antimicrobial resistance, epidemiology, outbreaks, phylogenomics

## Abstract

1.

**Background:** Drug-resistant tuberculosis is a high priority threat to global public health. There are still critical gaps in understanding how novel drug-resistant *M. tuberculosis* strains emerge and, once emergent, what drives the differential propagation of certain epidemiologically-successful strains over others. This study sought to describe the joint evolutionary and epidemiological histories of a novel multidrug-resistant *M. tuberculosis* strain recently identified in the capital city of the Republic of Moldova (MDR Ural/4.2).

**Methods:** Using whole genome sequence data and Bayesian phylogenomic methods, we reconstruct the stepwise acquisition of drug-resistance mutations in the MDR Ural/4.2 strain, estimate its historical bacterial population size over time, and infer the migration history of this strain between Eastern European countries.

**Results:** We infer that MDR Ural/4.2 likely evolved (via acquisition of *rpoB* S450L, which confers resistance to rifampin) in the early 1990s, during a period of social turmoil following Moldovan independence from the Soviet Union. This strain subsequently underwent substantial population size expansion in the early 2000s, at a time when national guidelines encouraged in hospital treatment of TB patients. We infer exportation of this strain and its INH-resistant ancestral precursor from Moldova to neighboring countries starting as early as 1985.

**Conclusions:** Our findings underscore how public health practice and social determinants of health shape the conditions under which *M. tuberculosis* evolves, and demonstrates how historical changes in these conditions shape present-day challenges in TB control. These findings underscore the need for regional coordination in TB control across Eastern Europe.

## 3. BACKGROUND

Drug-resistant tuberculosis (TB) remains an important priority in global public health (1). Drug-resistant tuberculosis, including multidrug-resistant (MDR) and extensively drug-resistant (XDR) forms, are highly endemic in Eastern Europe and countries of the Former Soviet Union (FSU). In Moldova, which records the highest per-capita rates of MDR-TB worldwide (54 cases per 100 000), nearly 25% of MDR-TB occurs among individuals without previous history of TB therapy and >50% of previously treated TB cases are MDR (2).

Drug-resistant TB cases in most high burden countries are attributable to ongoing transmission of specific, epidemiologically successful *Mycobacterium tuberculosis* (*M. tuberculosis*) strains, rather than *de novo* acquisition of drug resistance mutations while on anti-TB therapy (3-5). A large body of existing research has examined how social and economic turmoil, health systems failures, and mass incarceration following the collapse of the Soviet Union likely facilitated both selection for and transmission of drug-resistant TB in Eastern Europe (6). However, direct evidence linking specific historical events or public health practices to the emergence of drug-resistant *M. tuberculosis* strains is limited, particularly in heavily impacted countries like Moldova. This study reconstructs the evolutionary history of a recently identified (7), highly successful strain of MDR-TB currently circulating in Chisinau, Moldova (referred to here as *Ural/4*.*2*).

In this report, we employ whole genome sequencing (WGS) data to examine the genomic epidemiology of *M. tuberculosis* among a cross-sectional survey of TB patients diagnosed at a municipal hospital in Chisinau, Moldova. We find a high prevalence of MDR-TB and uncover the local predominance of a unique *M. tuberculosis* strain responsible for >60% of MDR-TB cases in this sample. Using phylodynamic methods, we examine the joint evolutionary and epidemiologic histories of this strain in Moldova and surrounding countries, yielding new evidence on how specific public health practices facilitated the emergence and subsequent dispersal of this important public health threat. These results support the expanded use of genomic surveillance in local and regional TB control efforts.

## 4. METHODS

### 4.1 Study site and participants

The study population included 404 individuals with culture-positive TB at the time of initiation of inpatient treatment at a municipal TB hospital in Chisinau, Republic of Moldova from October 2013 to December 2014. This TB hospital serves the municipality of Chisinau (population approximately 700,000). Male and female TB patients five years of age or over, both HIV negative and positive, were eligible for the study if they were able to provide informed consent and had a minimum of one positive *M. tuberculosis* culture result. Patients were excluded from the study if the quantity of respiratory secretions provided was less than 5ml, if their *M. tuberculosis* culture was negative, or if informed consent not provided. Viable serial isolates were processed for whole genomic sequence analysis using methods described below. Additional information on the study site and participant metadata are included in the Supplemental Information.

### 4.2 Ethical review

The study protocol was approved by the Institutional Ethical Committee for Research of Phthisiopneumology Institute, Chisinau, Republic of Moldova, Yale School of Public Health, and Columbia University Medical Center.

### 4.3 Genomic characterization of *M. tuberculosis* clinical isolates from the Republic of Moldova

Of 404 patient isolates collected in Chisinau, Moldova and sequenced via paired-end sequencing on the Illumina MiSeq platform, we recovered 376 (93%) *M. tuberculosis* isolates with WGS data meeting criteria for read depth (average read depth > 15 reads) and sequence coverage (reads covering > 99% of the reference genome). For phylogenetic analyses, we removed duplicate (serial) isolates by selecting, for each participant, the isolate with the earliest collection data, yielding a subset of n=283 non-duplicate individual isolates collected in Chisinau for analysis. Additional information on read alignment, filtering, and variant detection is included in the Supplemental Information.

### 4.4 WGS data for Eastern European and lineage-representative *M. tuberculosis* isolates

To provide regional context for the isolates collected in Chisinau, we obtained raw paired-end read data for n= 1168 samples included in the NCBI Sequence Read Archive, including those from prior regional studies of *M. tuberculosis* genomic diversity (8) and well-characterized isolates from relevant *M. tuberculosis* phylogeographic lineages to anchor important clades in the phylogenetic analysis. Read filtering, alignment, and variant calling and filtering for these additional sequences were conducted following the procedures described above.

### 4.5 Phylogenetic analysis

We conducted phylogenetic analysis on three groups of *M. tuberculosis* isolates, doing model testing in BEAST2(9), phylogenetic reconstruction and discrete trait analysis in BEAST 1.10 (10), and the *IQtree and timetree* tools available through Nextstrain *Augur* version 6.2.0 (11) (Supplemental Methods and **Supplemental Table E1**).

## 5. RESULTS

### 5.1 High prevalence of MDR-TB in the Republic of Moldova

Our cross-sectional study of *M. tuberculosis* genome sequences collected from October 2013 – December 2014 in Chisinau, the Republic of Moldova included yielded 283 patient isolates, representing approximately one quarter of all incident culture-positive TB cases diagnosed in Moldova during the same time period. A high proportion of isolates in this sample carried mutations associated antimicrobial drug resistance **(Figure 1A)**. 142 (50.2%) of these isolates carried at least one mutation associated with isoniazid resistance, 132 (46.7%) isolates were MDR by genotype (i.e. carried mutations associated with both isoniazid and rifampin resistance), and 45 (15.9%) of all isolates were MDR by genotype and carried *gyrA* mutations associated with resistance to fluoroquinolones (“pre-extensively drug resistant”, pre-XDR). Previous TB treatment (63% vs. 24%, *P<0*.*01*), history of homelessness (24% vs 14%, *P<0*.*05*), and urban residence (68% vs 54%, *P<0*.*05*) were factors associated with MDR or XDR-TB strains. (Here we apply the pre-2021 definition of XDR-TB, i.e. *M. tuberculosis* resistant to isoniazid and rifampin as well as fluoroquinolones and at least one injectable medication, such as streptomycin.) Among the 17 TB cases with HIV infection, 13 were MDR or XDR.

**Figure 1.**
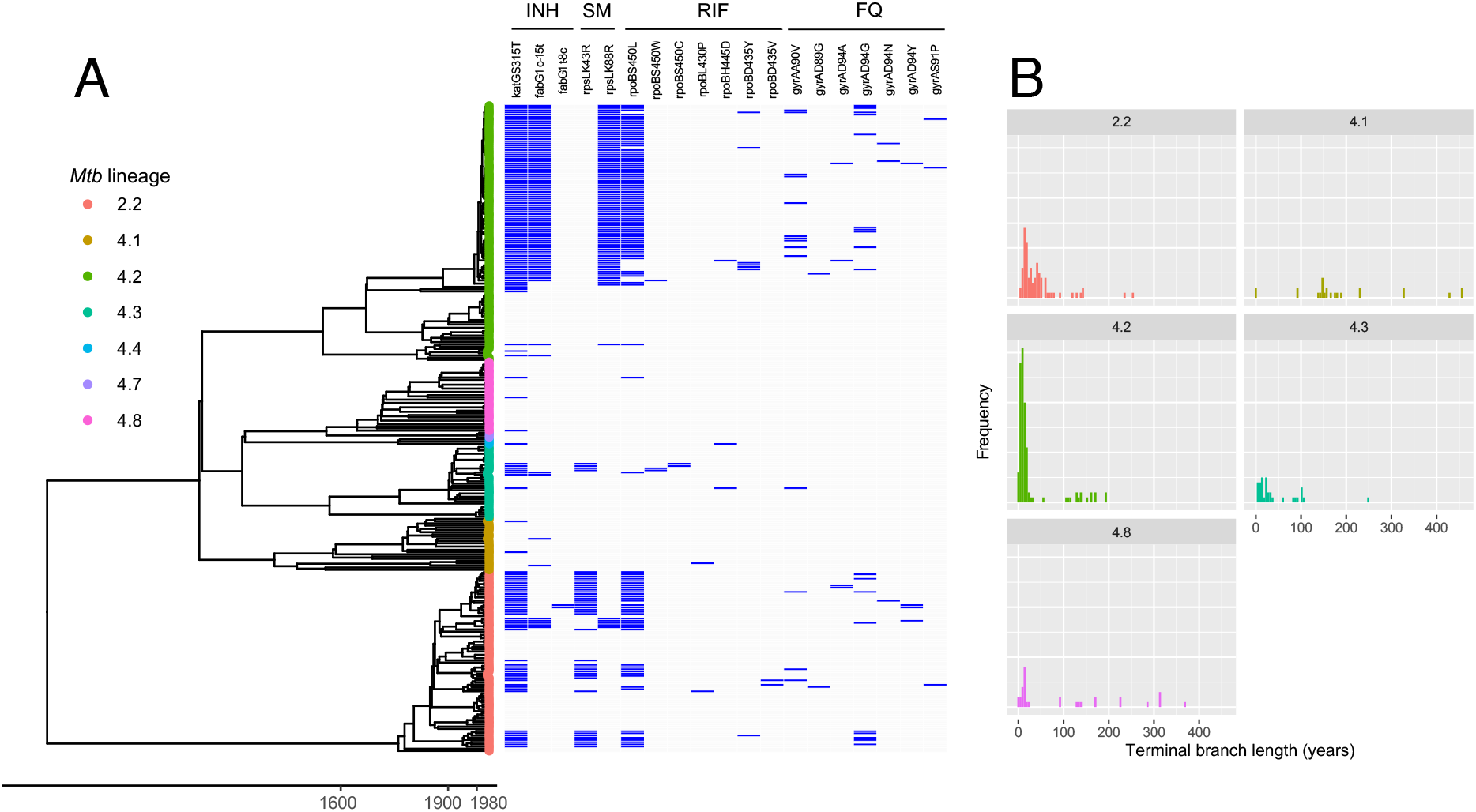
Bayesian phylogenetic analysis for n=293 *M. tuberculosis* isolates collected in the Republic of Moldova in 2013-2014. A: Dated phylogenetic reconstruction for all n=293 sequences collected in Chisinau and included in phylogenetic analysis. Tips of the tree are annotated by SNP barcode-based sublineage assignment (13). Selected mutations associated with antimicrobial resistance (specifically those associated with resistance to isoniazid, streptomycin, rifampin, and fluoroquinolones) are annotated in the adjacent heatmap. B: Distribution of terminal branch lengths (in years) by sublineage. Lineages 4.4 and 4.7 are not included in panel B given the small number of sequences in each clade. The phylogenetic tree was estimated using BEAST v1.10 with the HKY nucleotide substitution model, constant-size coalescent population model, strict molecular clock, and an informative prior on the mutation rate **(Supplemental Table E1)**.

### 5.2 MDR-TB in Moldova is diverse but dominated by a single epidemic strain

We next sought to understand the population structure of *M. tuberculosis* in our sample, and more specifically for isolates with MDR phenotypes. *M. tuberculosis* isolates collected during our study in Chisinau are comprised primarily of two major phylogenetic lineages of *M. tuberculosis*, Lineage 2 (L2) and Lineage 4 (L4) **(Figure 1A)**. There were 80 (28.3%) strains from phylogeographic lineage 2 and 203 (71.7%) strains from lineage 4. Using the SNP-based classification system described by Coll *et al*.,(13) all Lineage 2 strains were identified as belonging to sublineage 2.2.1; six different sublineages comprised the Lineage 4 isolates in our sample **(Figure 1A)**. The majority of all lineage 4 isolates in this sample are from sublineage 4.2 (n=113, 39.9% of all 283 isolates and 55.6% of L4 isolates).

Among the 132 genotypic MDR-TB isolates (including pre-XDR isolates), the majority (n=82, 62.1%) belong to the genetically monomorphic Ural/4.2 clade (**Figure 1A**). Using an adapted application of the Kolmogorov-Smirnov test that accounts for differences in the number of taxa in each clade (**Supplemental Figure E1**), we found that terminal branch lengths were significantly shorter for Ural/4.2 isolates when compared to Chisinau isolates in Lineages 2.2 and 4.1. These results consistent with recent and/or more frequent transmission of a single highly monomorphic strain (**Figure 1B, Supplemental Figure E1**), although this analysis is potentially subject to sampling bias. Terminal branch lengths for Ural/4.2 isolates were numerically smaller than those for lineage 4.3 and 4.8 isolates, but this difference was not statistically significant, potentially due to the smaller number of isolates from the latter two lineages. Patients harboring Ural/4.2 strains were more likely to have reported previous TB treatment when compared to other non-Ural/4.2 (62% vs 37%, *P<0*.*01*).

Among 77 total participants from Chisinau for which there is more than one isolate with available WGS data, we identified 8 participants who had discordant drug susceptibility genotypic profiles between serial isolates **(Supplemental Figure E2)**. Five of these participants had serial isolates with discordant lineage assignments and large SNP differences compared to their initial isolates **(Supplementary Figure E2, A-E)**, suggestive of either secondary infection with a different strain of *M. tuberculosis* following initial culture collection or an initial polyclonal infection with subsequent selection for the more drug-resistant isolate during anti-TB therapy. Among these four participants, MDR Ural/4.2 *M. tuberculosis* strains were recovered as the later, discordant isolate in two individuals. Three other participants **(Supplementary Figure E2, F-H)** had genotypic drug susceptibility profiles and inter-isolate SNP differences consistent with *de novo* acquisition of new drug resistance mutations between serial isolates. Of the remaining 69 participants with more than one isolate and concordant genotypic drug susceptibility profiles, none had discordant SNP-based lineage assignments, i.e. there was no evidence of polyclonal or secondary infection among participants with consistent drug susceptibility profiles across serial isolates.

### 5.3 Substantial proportions of incident TB cases are due to clustered transmission

We next assessed the extent of genomic clustering within our dataset, using two separate metrics: (1) SNP threshold, where isolates diverging by no more than 10 SNPs (with SNP differences calculated using all SNPs, including those associated with drug resistance) were considered clustered; and (2) a probabilistic approach that incorporates the SNP differences and information about case timing, the molecular clock rate, and transmission processes (12). Both methods were similar, indicating 36.2% and 36.9% were clustered by the cutoff and probabilistic approach, respectively. Previous TB treatment was associated (54% vs. 37%, *P<0*.*01*) with clustering by both methods.

### 5.4 Ural/4.2 strains are found across Eastern Europe

To place our analyses in a broader phylogenetic and geographic context, we combined sequence data from n=283 unique patient isolates collected in Chisinau with a larger set of publicly available *M. tuberculosis* genomes (N=1168) from Eastern Europe **(Figure 2)**. Isolates with genotypic multidrug resistance were widely prevalent in this collection, comprising 63.5% of all isolates in the dataset (**Figure 2**); 32.5% of all isolates (n=371) carry a *gyrA* mutation associated with fluoroquinolone resistance. *M. tuberculosis* clades generally distributed across multiple countries, with the exception of 4.2/Ural, which is largely comprised of Moldovan isolates collected during our study. 335 of 1451 isolates (23.1%) in this collection were identified as lineage 4.2/Ural, of which 275 (82.1%) were collected in Moldova. Excluding samples collected during our study in Chisinau, Ural/4.2 isolates comprise 14.7% (172 of 1168) of the larger collection of isolates from Eastern Europe. 112 of the 172 (65.1%) Ural/4.2 isolates from the regional collection of samples were collected in Moldova.

**Figure 2.**
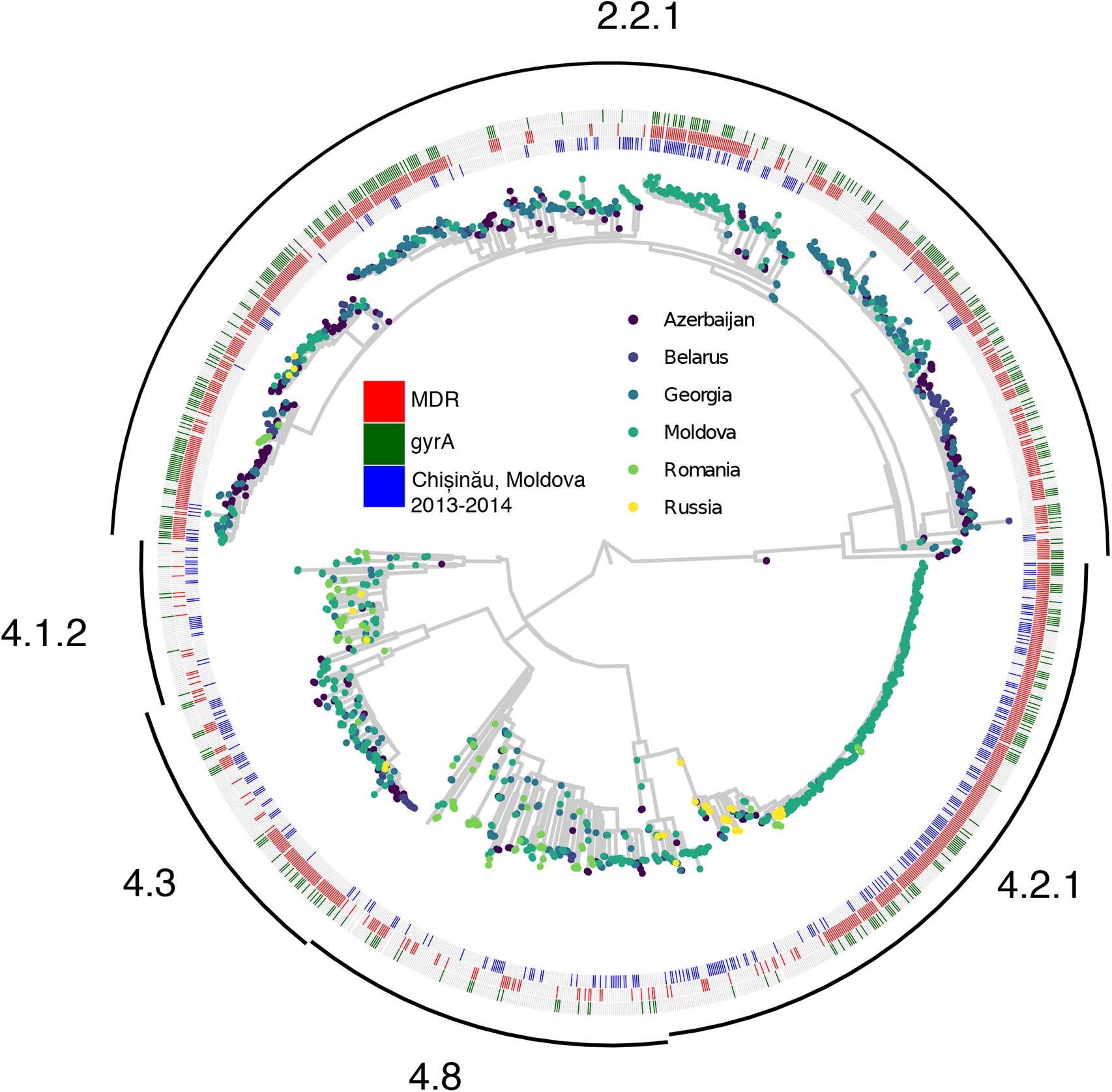
Bayesian phylogenetic reconstruction for n= 1451 *M. tuberculosis* isolates from Eastern Europe. Tips of the phylogeny are colored by the country of origin for each isolate. Annotations denote *M. tuberculosis* sequences collected in Chisinau during this study (inner ring, red) versus sequences from prior studies (inner ring, grey), sequences with genotypic multidrug-resistance (middle ring, blue), and sequences with any fluoroquinolone resistance-associated *gyrA* polymorphism (outer ring, blue). The phylogenetic tree was estimated using the *IQtree* and *timetree* tools in Nextstrain *Augur* 6.2.0.(11) Additional details on this group of samples is available in **Supplemental Table E1**. Clades of the tree are annotated with SNP barcode-based sublineage assignments.(13)

### 5.5 Phylogenetic reconstruction indicates emergence of MDR Ural/4.2

To examine the timeline over which the Ural/4.2 outbreak strain acquired multidrug resistance, we estimated time to most recent common ancestor (TMRCA) for clades carrying canonical drug resistance mutations in *inhA* (including its associated promoter region) and *katG* (associated with resistance to isoniazid), *rpoB* (associated with resistance to rifampin), and *gyrA* (associated with resistance to fluoroquinolones) on the phylogenetic tree (**Figure 3**). We define the *emergence* date for MDR Ural/4.2 as the TMCRA isolates carrying the strain-specific *rpoB* mutation S450L, which confers resistance to rifampin and the MDR phenotype. We note that undetected homoplasic acquisition of drug resistance mutations within clades can potentially bias these estimates toward longer TMRCA (earlier dates of emergence) and, where this possibility is suggested by the phylogenetic tree, we report ages for multiple nodes representing potential most recent common ancestors for a given mutation **(Figure 3A)**. We estimated that Ural/4.2 evolved resistance to isoniazid (via acquisition of *katG* S315T) in approximately 1984 (Node A, 95%HPD: 1970-1995) and acquired the *rpoB* S450L mutation, conferring resistance to rifampin, at some time between 1983-2000 (Node J, TMRCA: 1993). A candidate compensatory mutation in *rpoC* V483G arose coincident with *rpoB* S450L (**Supplementary Figure S3)**. Additional Ural/4.2 mutations in *rpsL* K88R (conferring resistance to streptomycin) and *inhA* promoter -C15T (conferring resistance to isoniazid) were acquired in 1984 (Node A, 95%HPD: 1970-1995) and 1987 (Node B, 95% HPD: 1976-1997), respectively. A variety of *gyrA* mutations are observed across Ural/4.2 isolates, suggesting that MDR-TB Ural/4.2 strains independently acquired *gyrA* mutations multiple times, subsequent to the acquisition of mutations associated with isoniazid and rifampin resistance. Thus, phylogenetic analysis suggests MDR Ural/4.2 strains emerged in the early 1990s acquiring multiple resistance mutations (i.e. in *katG, inhA promoter, rpoB*) prior to expansion as a highly drug resistant clone. We found no known resistance-associated polymorphisms in genes previously implicated in resistance to bedaquiline (*Rv0678, atpE*, and *pepQ(14)*) among isolates from Chisinau.

**Figure 3.**
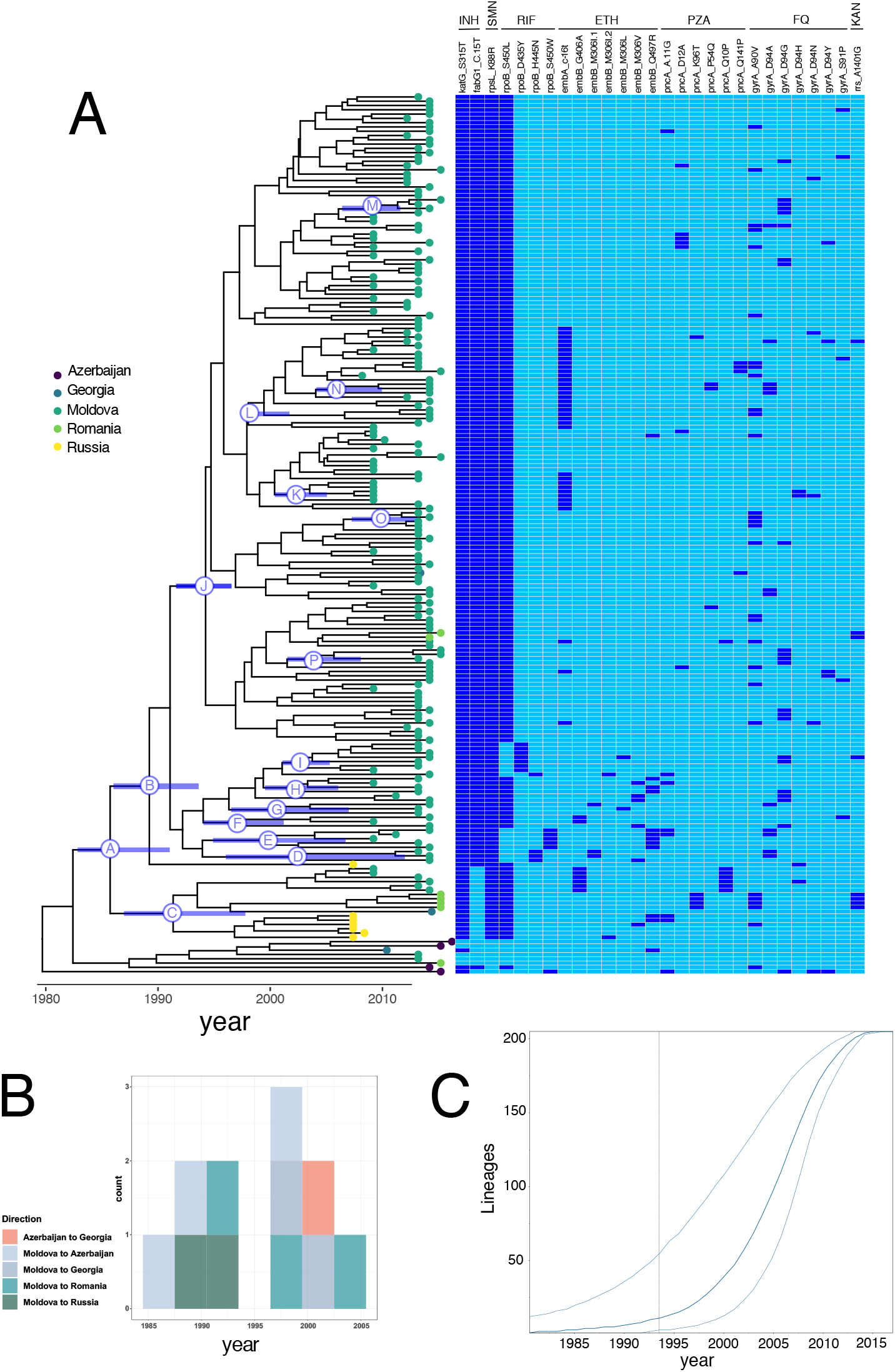
Bayesian phylogenetic reconstruction with discrete trait analysis for country of origin for n=205 *M. tuberculosis* isolates from the Ural/4.2.1 lineage, including the multidrug resistant outbreak strain and closely related isolates. A: Phylogenetic reconstruction and estimated TMRCA for isolates carrying known *M. tuberculosis* drug resistance mutations. Nodes corresponding to most recent common ancestor for relevant drug resistance mutations are labeled A-O, with the 95% highest probability density for node age displayed the blue bar on each labeled node. Tips are labeled with the country of origin for each isolate (Azerbaijan, Georgia, Moldova, Romania, and Russia). Median node ages and 95%HPD (listed in parentheses are as follows: A: *katG* S315T 1984 (1970-1995); B: *fabG* C(15)T 1987 (1976-1997); C: *rpoB* S450L 1990 (1979-1999); D: *rpoB* H455N 2001 (1992-2009); E: *rpoB* S450W 1998 (1990-2005) F: *rpoB* S450L 1995 (1986-2003); G: *rpoB* S450L 1999 (1991-2007); H: *rpoB* S450L 2011 (1995-2006); I: *rpoB* D435Y 2001 (1996-2006); J: *rpoB* S450L 1993 (1983-2000); K: *embA* C(16T) 2001 (1996-2006) L: *embA* C(16T) 1996 (1989-2003); M: *gyrA* D94G 2008 (2005-2011); N: *gyrA* D94A 2005 (1999-2010; O: *gyrA* A90V 2009 (2005-2011). B: Discrete trait analysis for migration events. Boxes show counts for estimated number of migration events between countries by year and are colored according to pairs of sending and receiving countries. C: Estimated effective population size over time with exponential demographic model for Bayesian phylogenetic reconstruction. Outer lines show the 95% highest posterior density interval for the effective population size.

**Figure 3C** shows the estimated *M. tuberculosis* bacterial population size for Ural/4.2 isolates over time inferred from Bayesian phylogenetic reconstruction using an exponential demographic model (the best-supported model in our analysis, see supplemental methods). Using this model, we observe substantial expansion of the *M. tuberculosis* effective population size (*Ne*) starting between 1995 and 2000 **(Figure 3C)**. To further test the hypothesis that Ural/4.2 *M. tuberculosis* bacterial population size expanded recently, we used non-parametric Bayesian skyline analysis to estimate the historical size of Ural/4.2 population. Consistent with the exponential demographic model, we observed expansion in the effective population size of Ural/4.2 strains starting on or around the year 2000 **(Supplemental Figure E4)**. Model testing for Bayesian phylogenetic analysis, which favors exponential growth as the best-performing demographic model and the aforementioned Bayesian Skyline as the second-best model **(Supplemental Table E2)**, provides additional supporting evidence for recent prior population growth, consistent with relative increases in overall transmission, for the Ural/4.2 outbreak clade. Taken together, our results indicate that MDR Ural/4.2 clade emerged in the early 1990s achieving sustained transmission in the early 2000s with resultant increases in bacterial population size. At the time of this study, almost all TB patients initiated treatment as inpatients, and average length of hospitalization exceeded two months as recently as 2016.

### 5.6 Geographic origin and regional dispersal of MDR Ural/4.2 strains

Next, we performed analyses to formally examine the phylogeographic history of Ural/4.2 strains in Eastern Europe treating sampling location (country) as a discrete trait. Applying DTA, the geographic location of the MRCA for Ural/4.2 sequences was estimated to be Moldova with posterior probability = 0.66, although this inference may be influenced by unsampled locations or regions **(Figure 3B)**. To investigate the migration history of Ural/4.2 over time, we analyzed the inferred load and direction of migration over time (see Supplemental Information). Inferred migration events of Ural/4.2 strains indicate drug-resistant and MDR Ural/4.2 strains were exported from Moldova to neighboring countries (Azerbaijan, Georgia Romania, Russia) from mid-1980s onwards, with additional estimated migration events between Azerbaijan and Georgia after the year 2000.

## 6. DISCUSSION

Countries in Eastern Europe record the highest incidence rates of drug resistant tuberculosis globally. Here, we report transmission of a specific, epidemiologically successful strain of *M. tuberculosis* (MDR Ural/4.2) as the most frequent cause of MDR-TB in a cross-sectional cohort of patients in Chisinau, Moldova. Our results indicate that this strain evolved in the early 1990s and underwent marked population expansion after achieving sustained transmission in the early 2000s. Crudu et al previously reported that nosocomial re-infection resulting in MDR-TB in the Republic of Moldova was often attributable to a MDR-TB Ural/4.2 strain (15). Since this study focused only on Chisinau, we do not yet know the extent to which this strain circulates within the rest of Moldova. However, we do find limited phylogenetic evidence indicating recent exportation of MDR Ural/4.2 from Moldova to neighboring countries in Eastern Europe, underscoring the need for regional coordination in TB surveillance and control.

There is increasing evidence that transmission of existing drug-resistant strains is the primary driver of new infections in most drug-resistant TB epidemics (3, 5). We find that despite accounting for only a small proportion of the drug susceptible bacillary population, Ural/4.2 strains are heavily enriched among MDR and pre-XDR groups of isolates. Short terminal branch lengths indicate that this enrichment is most likely driven by recent or more frequent transmission. (Alternatively, shorter branch lengths observed in the Ural/4.2 clade could be attributable geographic or other kinds of sampling bias, if such bias resulted in preferential sampling of clustered Ural/4.2 isolates and/or under-sampling of isolates from other clades, but it is difficult to measure or account for this hypothetical source of bias.)

Consistent with recent findings from Sinkov et al. (7), we find that MDR-TB Ural/4.2 strains emerged in the early 1990s, approximately at the time of Moldovan independence from the Soviet Union. This period was characterized by a decade of economic turmoil, mass incarceration, underfunded public health programs, and unreliable TB drug supplies (16). This complex interplay between socio-economic conditions and poor TB case management likely created conditions conducive to the evolution of MDR and other drug-resistant *M. tuberculosis* strains.

We infer from Bayesian phylogenetic analysis that Ural/4.2 has undergone substantial and relatively recent *M. tuberculosis* bacterial population expansion. Using both an exponential demographic model and the non-parametric Bayesian Skyline method (two of the best-performing demographic models in our analysis per marginal likelihood estimation), we observe evidence of substantial increases in the Ural/4.2 bacterial population from the year 2000 onward. During this time period, as per Moldovan TB control policy, all TB patients were hospitalized during the intensive phase of therapy and cohorting of MDR and non-MDR patients occurring only after phenotypic DST results were available (17). We find that MDR-TB Ural/4.2 strains were more likely recovered from individuals with previous TB history and that Ural/4.2 strains were isolated from approximately half of all follow-up discordant serial isolates, suggesting a role for exogenous reinfection during anti-TB therapy. This phenomenon, i.e. secondary or polyclonal infection identified via genotyping of serial isolates, is observed in other cross-sectional *M. tuberculosis* genomic surveillance studies where an epidemiologically-successful drug-resistant strain is circulating in the community (19, 20).

There is concern that MDR and pre-XDR *M. tuberculosis* strains can disseminate beyond international borders and seed neighboring and more distant countries (21, 22). Our analysis indicates that Ural/4.2 was exported from Moldova to at least four countries in Eastern Europe between ∼1985 and 2005 **(Figure 3C)**. Although we document multiple cross-border strain migration events, extensive regional dissemination appears to be limited, consistent with prior studies on the global distribution of drug-resistant TB strains (23). We acknowledge that our collection of 4.2/Ural isolates is prominently over-sampled for isolates collected in Chisinau and under-sampled for isolates from the rest of the country and from countries other than Moldova, and that this is an important limitation on our ability to infer historical migration events from the available data, although expect that this limitation would result in bias toward inferring fewer migration events overall. Our findings indicate regional dissemination may be nascent and aggressive approaches to contain MDR Ural/4.2 strains locally may prevent further cross border events.

## 7. CONCLUSIONS

Taken together, our results underscore the intersection of socio-economic factors and public health policy in shaping drug resistant TB epidemics. Our findings highlight how large systematic shortcomings in clinical, public health and social programs can facilitate the emergence and transmission of drug-resistant TB. Extensive nosocomial transmission was the key catalyst for the subsequent rapid population expansion of Ural/4.2 in Moldova. To avoid widespread regional transmission of highly drug resistant strains, adequate resources for local containment and regional control efforts will be required. Our study supports regional cooperation and coordination in TB control (24, 25) and underscores the need for regional public access data that systematically incorporates epidemiological and pathogen information.

## Data Availability

Whole genome sequencing data for isolates collected in Chişinău, Moldova are available in the National Center for Biotechnology Information Sequence Read Archive (NCBI SRA bioproject number: PRJNA482716). Whole genome sequence data for the regional collection of isolates from eastern Europe are available under NCBI SRA bioproject numbers PRJNA421446, PRJNA429460, PRJNA384815, PRJNA318002, PRJNA480888, and PRJNA482865.

## 8. LIST OF ABBREVIATIONS

MDR: multidrug-resistant;
TB: tuberculosis;
SNP: single nucleotide polymorphism;
WGS: whole genome sequence;
XDR: extensively drug-resistant

## 9. DECLARATIONS

### Ethics approval and consent to participate

The study protocol was approved by the Institutional Ethical Committee for Research of Phthisiopneumology Institute, Chisinau, Republic of Moldova (Approval Oct 2012), Partners Human Research Committee (2012-P-001904/1), Yale University Human Investigation Committee (IRB number: 1409014628), and Columbia University Medical Center (Not Human Subjects Research Under 45 CFR 46).

### Availability of data and materials

Whole genome sequencing data for isolates collected in Chisinau, Moldova are available in the National Center for Biotechnology Information Sequence Read Archive (NCBI SRA bioproject number: PRJNA482716). Whole genome sequence data for the regional collection of isolates from eastern Europe are available under NCBI SRA bioproject numbers PRJNA421446, PRJNA429460, PRJNA384815, PRJNA318002, PRJNA480888, and PRJNA482865.

### Authors’ contributions

TSB., VE, OB, and MO planned and conducted the data analysis and co-wrote the manuscript. NS planned and conducted data analysis. JS developed analytical methods used for strain clustering. CC supervised data analysis and co-wrote the manuscript. SA, EN, and NC supervised and conducted laboratory analysis. VC, THC, and BM designed the study, supervised laboratory work and data analysis, and co-wrote the manuscript.

## Acknowledgements

We would like to acknowledge in kind support from the American Museum of Natural History for providing computational resources.

