## Supplemental_Information for "Evolution and emergence of multidrug-resistant *Mycobacterium tuberculosis* in Chisinau, Moldova"

### 1. SUPPLEMENTAL METHODS

#### 1.1 Study site and additional data

At the time of this study, the municipal hospital in Chișinău, the Republic of Moldova (the study site) had approximately 375 inpatient beds, 50 of which were typically reserved for MDR-TB patients, and served over 1220 hospitalized patients per year. The catchment area for this hospital includes the city of Chișinău and neighboring localities. All epidemiologic and laboratory data from TB patients are routinely entered into a country-wide web-based electronic surveillance system for monitoring and evaluation of TB control programming (SIMETB, https://simetb.ifp.md/). Epidemiologic data including age, sex, previous TB history, results of chest radiograph, history of incarceration, and place of residence were collected. Laboratory data, including mycobacterial smear grade, culture and drug-susceptibility testing to first and second line antituberculosis agents, were extracted from the electronic medical record.

#### 1.2 Mycobacterial culture and drug susceptibility testing

Lowenstein-Jensen (LJ) culture was performed from sputum specimens using standard NALC-NaOH decontamination. All sputum cultures positive for MTB complex subspecies tuberculosis were subjected to first-line drug susceptibility testing (DST) for isoniazid (critical concentration of 1 µg/mL and 0.2 µg/mL), rifampicin (40 µg/mL), streptomycin (4 µg/mL) and ethambutol (2 µg/mL) using the absolute concentration method. Any resistant strains underwent further DST for second-line agents performed using the indirect proportion method on 7H11 agar as previously described.(1) DNA was extracted using the CTAB method.

#### 1.3 Whole genome sequence data processing

Paired-end (250 base pair) sequences were generated on the Illumina MiSeq platform. Raw paired-end reads were filtered for length and trimmed for quality (Trim Galore, Babraham Bioinformatics), removing read ends with Phred quality score < 15 and requiring overlap of at least 7 bp for removal of adapter sequences on each read, and duplicate reads were removed following alignment to the H37Rv reference genome (NC_000962.3) using the Burrows-Wheeler Aligner (2), similar to the pre-processing pipeline described by O'Neill et al (3). SNPs were identified using Samtools v0.1.19(4) and filtered for quality, read consensus (>75% reads supporting the alternate allele), and proximity to indels. Polymorphisms in or within 50 base pairs of hypervariable PPE/PE gene families, repeat regions, and mobile elements were excluded, similar to prior studies using WGS from *Mtb* (5, 6). We assigned each sequence in the study a SNP barcode-based sublineage assignment following Coll *et al* (7). Sequence data is publicly available under NCBI Sequence Read Archive accession number SRP156366. We typed drug resistance-associated variants from WGS data using Mykrobe Predictor (version 0.3.6(8)) with the "wheeler-2015" database of resistance variants. We applied the Kolmogorov-Smirnov test to evaluate whether terminal branch lengths differ between clades in a phylogenetic tree (where shorter branch lengths are considered a proxy for more frequent or more recent transmission of a given clade), and apply a modified version of this test to account for the different number of taxa in each clade (**Supplemental Figure E1**)(6).

#### 1.4 Metrics for genomic clustering

To measure genomic clustering, a measure of genetic relatedness used to infer transmission, we use two approaches. First, we approach is the widely used SNP threshold method: group(s) of strains differing by no more than 10 SNPs were assigned into a cluster. Second, we employ a probabilistic approach that considers absolute SNP differences and the length of time over which those differences have accumulated, using information about case timing (isolate collection date), the molecular clock of *Mtb*, and transmission processes.(9) We implement this method in R package *Transcluster* (https://github.com/JamesStimson/transcluster) (9).

**1.5 Bayesian phylogenetic reconstruction and discrete trait analysis**

*Model testing:* We used Markov Chain Monte Carlo (MCMC)-based Bayesian phylogenetic inference in BEAST2 (10) to reconstruct dated phylogenetic trees and infer the time to most recent common ancestor (TMRCA) for isolates carrying drug resistance mutations of interest. Model testing was done in BEAST 2 by means of nested sampling to calculate marginal likelihoods (10). Constant, exponential, skyline and extended skyline demographic models were combined with either a strict or a log normal relaxed clock. In all runs, *bModelTest* (11) which allows the MCMC to switch between substitution models, were employed rather than specifying a substitution model. **Supplemental Table E2** gives the marginal likelihood values for models tested. The resultant marginal likelihoods provided highest support for a nucleotide substitution model with transition-transversion parameters similar to the HKY-Γ, which was used in subsequent analyses.

*Phylogenetic inference with discrete trait analysis:* We used the best-performing model (HKY-Γ, relaxed molecular clock, exponential growth demographic model) in BEAST 1.10 with discrete trait analysis to infer phylogenies and migration events for the dataset of Ural/4.2 isolates (Group 3, **Supplemental Table E1**). Country of isolation (Moldova, Russia, Georgia, Azerbaijan, or Romania) was treated as a discrete trait with an asymmetric substitution model. Three independent chains of 250 million steps were assessed for convergence and combined. For the group of all samples collected in Chișinău during our study (Group 1, **Supplemental Table E1**), in which we sought to reconstruct phylogenetic relationships without detailed inference of other features (e.g. TMRCA or migration history) we used a simpler model with the HKY substitution model, constant-size coalescent population model, and an informative prior on the mutation rate (specifically, a normal distribution around 1E-7 mutations/site-year).

Given that the three best models (constant, exponential and skyline) with a relaxed clock were indistinguishable in terms of their marginal likelihood estimates (overlapping in their estimates ± 2 times the sum of their standard deviations, **Supplementary Table E2**), we ran subsequent analyses for the exponential demographic model, and the flexible skyline model (**Supplementary Figure E3**) that should also be able to capture constant tree dynamics fairly well. Employing the skyline model resulted in large highest posterior density intervals for the root TMRCA (1079-1868). Apart from the earliest branching events, the output from the skyline model was largely concordant with the output from the exponential model, with dense branching and diversification starting around 1995 and multiple exports inferred from Moldova between 1995 and 2005. The major difference between the outputs from the DTA analyses employing the exponential and skyline models were in the deepest branches of the phylogeny, where the skyline model inferred an origin in Azerbaijan with a few early export events to Georgia, Moldova and Romania, whereas the exponential model inferred a more recent origin in Moldova, followed mainly by exports from Moldova to other Eastern European countries.

*Temporal signal testing:* We tested the hypothesis that the sequence data under consideration was sufficient to inform estimates of both phylogenetic structure and molecular clock rates (“temporal signal testing”), two ways: (1) by evaluating the correlation, via linear regression, between root-to-tip distance and tip dates for a maximum likelihood phylogenetic tree reconstructed without information on tip dates, and (2) by randomizing tip dates between different sequences for n=10 independent BEAST runs and evaluating whether the clock rate and its associated uncertainty estimated using the non-randomized (true) tip dates is non-overlapping with estimates using the randomized tip dates. Root-to-tip regression was performed using Tempest(12), resulting in R0=0·096 and an adjusted p-value = 3·56E-06 for the tree constructed from sequences in the Ural/4.2 dataset. Randomizing tip dates across taxa yielded multiple BEAST runs in which the 95%HPD interval for estimated mutation rate overlaps with the 95%HPD interval for the BEAST run with non-randomized (true) tip dates, indicating absence of strong temporal signal data in the available sequence data (**Supplemental Figure E5**). Despite the weak evidence for temporal signal in our data, our analyses, which used a uniform prior for the molecular clock rate spanning 1E-10 to 1E-5, converged to estimated clock rates that were repeatedly within previously reported values (median: 1.7E-7, 95% HPD interval: 1.2366E-7-2.1673E-7), and thus we did not specify a more strongly-informative rate prior.

#### 2. SUPPLEMENTAL FIGURES

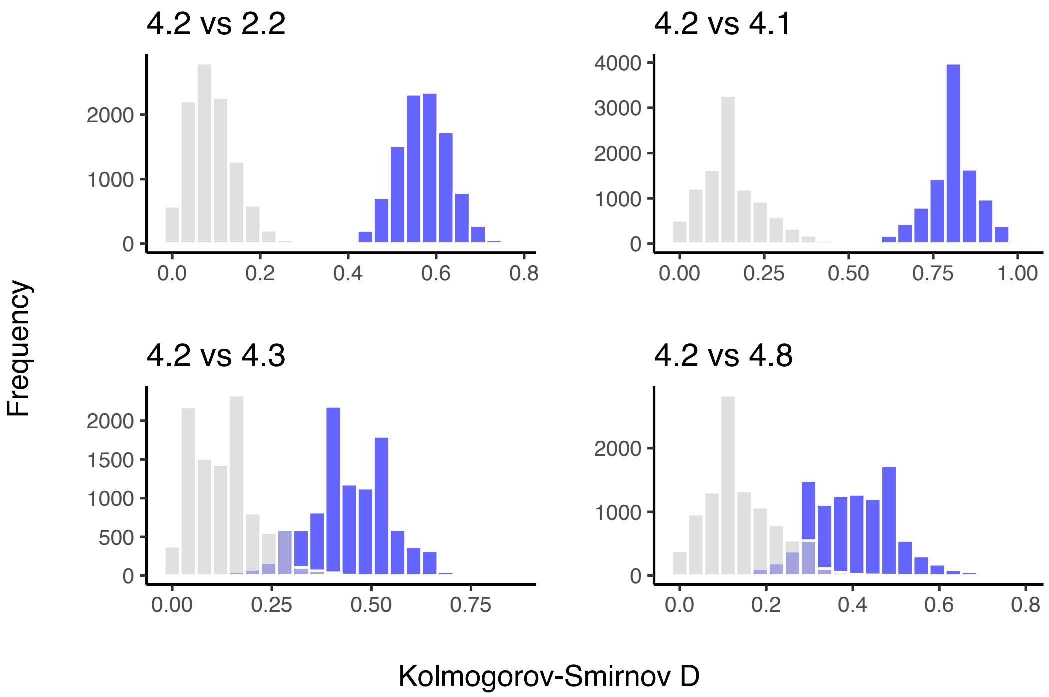


**Supplemental Figure E1**. Comparison between terminal branch lengths by clade. Observed (blue) and null (grey) distributions of Kolmogorov-Smirnov *D* values from equal-sized bootstrap samples from each clade. To calculate the distribution of observed *D* values, we select two evenly sized bootstrap samples from each clade of interest (taxa and corresponding terminal branch length selected randomly with replacement), with size equal to the number of taxa in the smaller of the two clades, calculate *D* (one-sided Kolmogorov-Smirnov testing whether values in 4.2 are smaller than those in each comparison clade), and repeat this procedure 10,000 times. Branch lengths are measured in years, estimated in BEAST 1.10 for the collection of isolates in Figure 1. To calculate the null distribution, we select evenly-sized bootstrap samples as above, but then randomize clade assignment across taxa before calculating *D*. Non-overlapping observed and null distributions support the alternate hypothesis that branches of clade 4.2, when accounting for the number of taxa in different clades under comparison, are significantly shorter than the comparator clade.

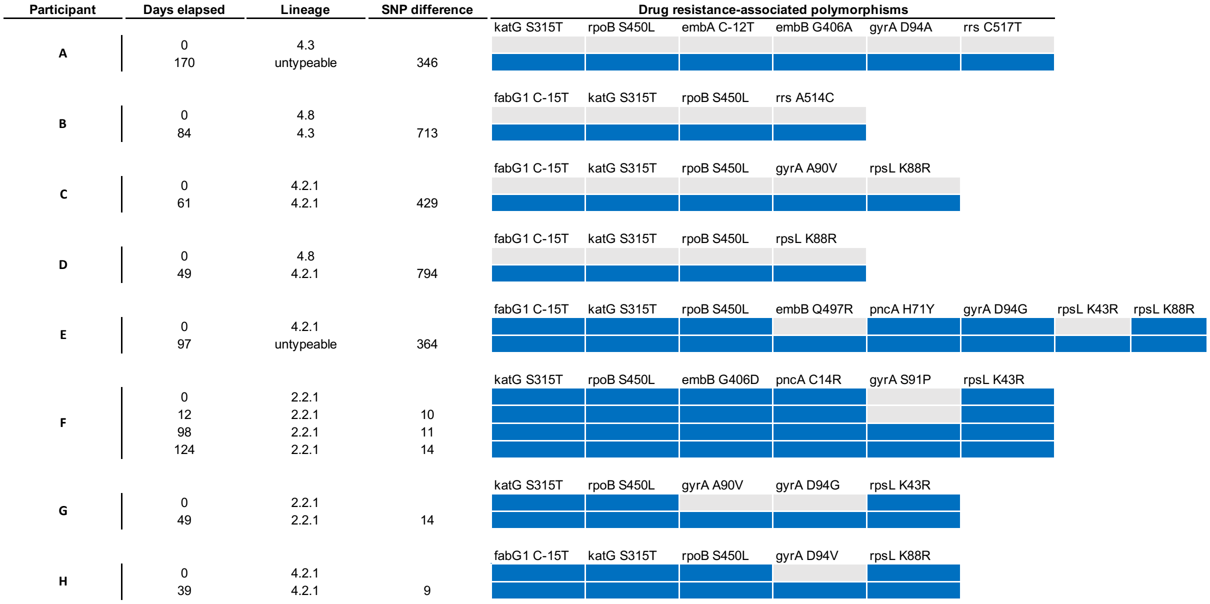


**Supplemental Figure E2**. Drug resistance polymorphisms for isolates from n=8 participants with discordant antibiotic susceptibility genotypic profiles between serial isolates. Days elapsed between culture collection dates, SNP-based lineage assignment, and number of SNP differences between isolates are listed for participants A-H. Two isolates with low average read depth could be given a SNP-based lineage and are thus listed as "untypeable". Participants A-E, characterized by discordant lineage assignments and relatively large SNP differences between isolates, likely represent secondary or initial polyclonal infection with a more highly drug-resistant strain. F-H, where lineage assignments are concordant between isolates and SNP differences between isolates are smaller, likely represent participants who acquired *de novo* drug resistance while on anti-TB therapy.

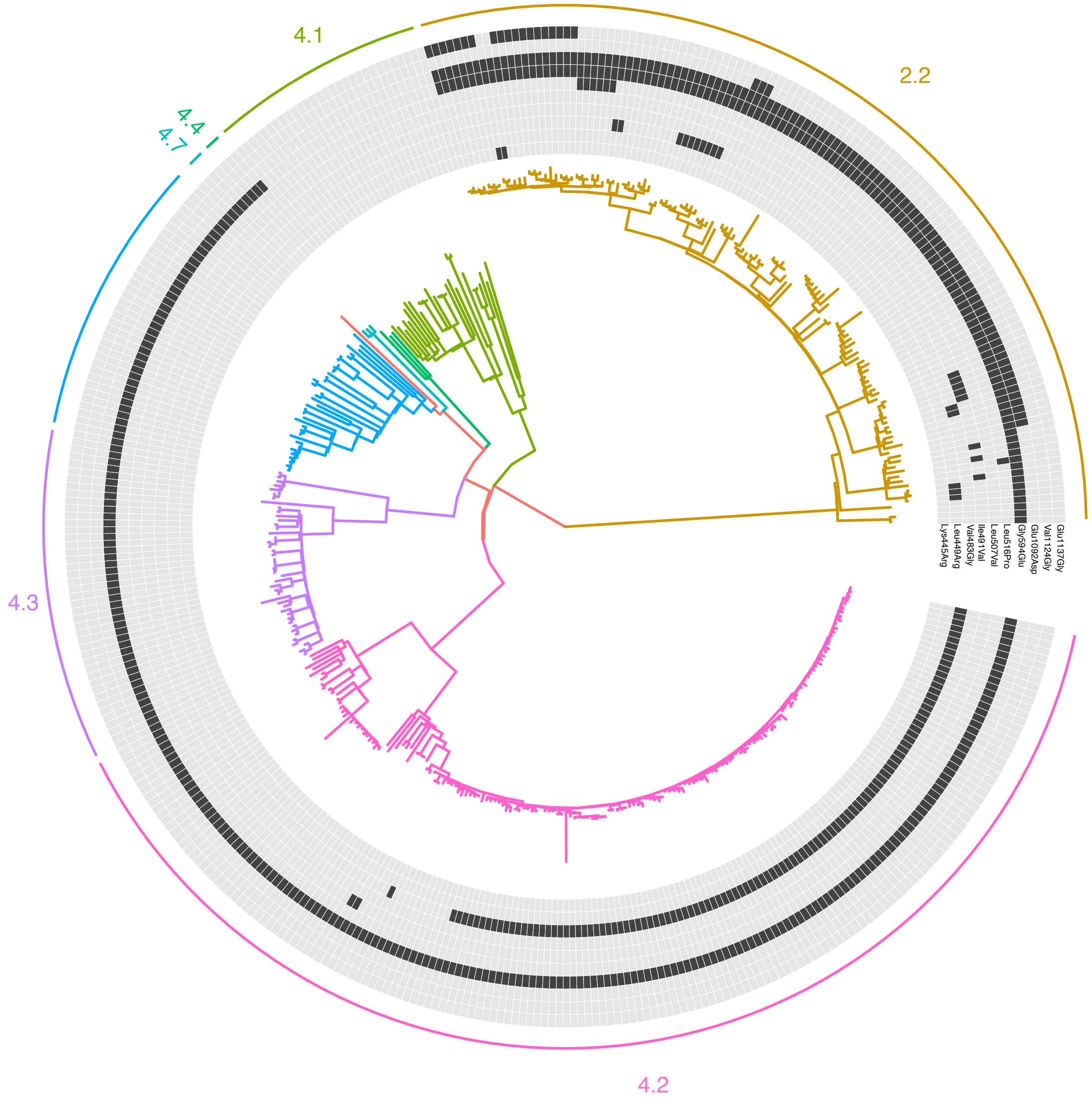


**Supplemental Figure E3. Non-synonymous *rpoC* mutations for isolates collected in Chișinău, Moldova.** Clades are colored and labeled with their respective phylogeographic lineages, assigned using the SNP barcode-based system in Coll *et al(7)*.

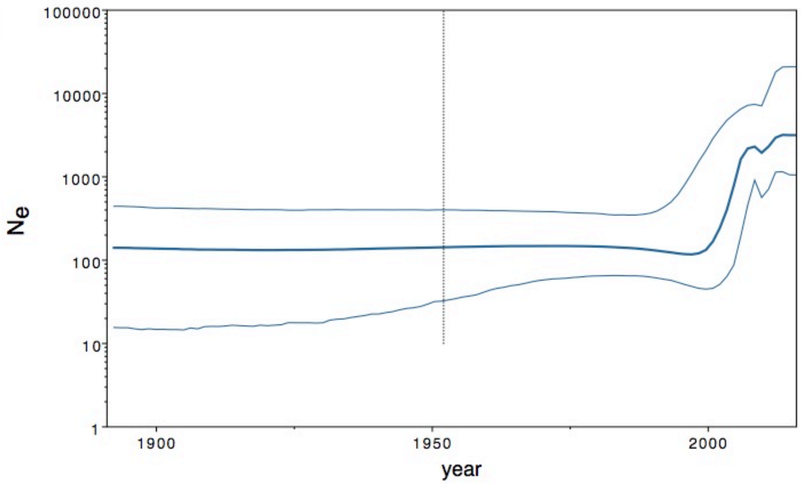


**Supplemental Figure E4. Estimated demographic history for the Ural/4.2 *Mtb* outbreak strain using Bayesian Skyline population model in BEAST**. Dark line shows median estimated effective population size ($N_e$) over time and thin lines demarcate the 95% highest probability density interval for $N_e$

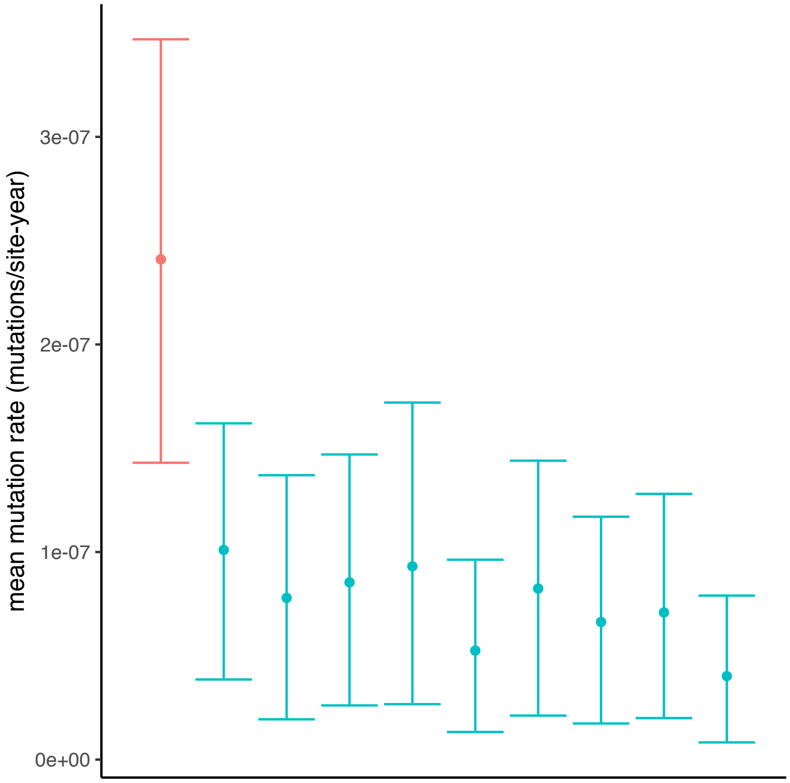


**Supplemental Figure E5 Temporal signal testing for n=205 sequences used for phylogenetic analysis of Ural/4.2 outbreak clade.** Red: Mean mutation rate with 95%HPD interval for BEAST analysis with non-randomized (true) tip dates. Green: Mean mutation rate with 95%HPD interval for BEAST analyses with randomized tip dates.

#### 3. SUPPLEMENTAL TABLES

| Group | Num. of isolates | Description | Methods for phylogenetic inference | Notes |
|---|---|---|---|---|
| 1 | 293 | Includes n=283 unique patient isolates collected in Chișinău, Moldova for this study plus n=10 representative archival isolates from selected *Mtb* phylogeographic lineages. | BEAST v1.10 | Implemented with HKY nucleotide substitution model, constant size coalescent population size, and strict molecular clock with an informative prior on the mutation rate. |
| 2 | 1451 | Includes Chișinău patient isolates plus WGS data for n=1168 isolates from Moldova, Russia, Georgia, Azerbaijan, and Romania included in the TB Portals database(13) | Nextstrain (Augur version 6.2.0) | Implemented using *IQtree* and *timetree(14)* |
| 3 | 205 | Includes Lineage 4.2/Ural isolates from Chișinău (collected in this study) plus representative 4.2 Ural isolates from neighboring countries. | BEAST v1.10 with discrete trait analysis | Implemented with HKY nucleotide substitution model, exponential coalescent population model (and Bayesian skyline plot in supplemental analysis), relaxed molecular clock, and uniform prior on the mutation rate. Model parameters were selected via nested sampling in *bmodeltest*.(11) |

**Supplemental Table E1.** Details on sample groups, their respective sample sizes and data sources, and the phylogenetic methods used for each group.

| Site model | Clock | Tree prior | Marginal likelihood | SD |
|---|---|---|---|---|
| Bmodeltest | Relaxed lognormal | Exponential | -13885.1488 | 8.3066 |
| Bmodeltest | Relaxed lognormal | Skyline | -13903.8632 | 8.4641 |
| Bmodeltest | Relaxed lognormal | Constant | -13911.9382 | 8.6872 |
| Bmodeltest | Relaxed lognormal | Extended skyline | -13927.5528 | 8.4392 |
| Bmodeltest | Strict | Skyline | -13948.8936 | 8.2061 |
| Bmodeltest | Strict | Exponential | -13965.9866 | 7.7838 |
| Bmodeltest | Strict | Constant | -14009.2728 | 7.9903 |
| Bmodeltest | Strict | Extended skyline | -14206.473 | 8.537 |

**Supplemental Table E2.** Model testing for Bayesian phylogenetic analysis in BEAST
